# Risk factors for SARS-CoV-2 infection among farmworkers in Monterey County, California

**DOI:** 10.1101/2021.02.01.21250963

**Authors:** Ana M. Mora, Joseph A. Lewnard, Katherine Kogut, Stephen A. Rauch, Norma Morga, Samantha Hernandez, Marcus P. Wong, Karen Huen, Cynthia Chang, Nicholas P. Jewell, Nina Holland, Eva Harris, Maximiliano Cuevas, Brenda Eskenazi, on behalf of the CHAMACOS-Project-19 Study Team

## Abstract

**Importance:** Essential workers in agriculture and food production have been severely affected by the ongoing COVID-19 pandemic.

**Objective:** To identify risk factors associated with SARS-CoV-2 shedding and antibody response in farmworkers in California.

**Design:** This cross-sectional study collected survey data and determined current SARS-CoV-2 shedding and seropositivity among 1,107 farmworkers in California’s Salinas Valley from 16 July to 30 November 2020.

**Setting:** Farmworkers receiving transcription-mediated amplification (TMA) tests for SARS-CoV-2 infection at federally qualified community clinics and community sites were invited to participate in our study.

**Participants:** Individuals were eligible if they were not pregnant, ≥18 years old, had conducted farm work since the pandemic started, and were proficient in English or Spanish.

**Exposures:** Sociodemographic, household, community, and workplace characteristics.

**Main Outcome(s) and Measure(s):** Current (as indicated by TMA positivity) and historical (as indicated by IgG seropositivity) SARS-CoV-2 infection.

**Results:** Most farmworkers enrolled in the study were born in Mexico, had primary school or lower levels of educational attainment, and were overweight or obese. Current SARS-CoV-2 shedding was associated in multivariable analyses with attained only primary or lower educational levels (RR=1.32; 95% CI: 0.99-1.76), speaking an indigenous language at home (RR=1.30; 0.97-1.73), working in the fields (RR=1.60; 1.03-2.50), and exposure to known or suspected COVID-19 case at home (RR=2.98; 2.06-4.32) or in the workplace (RR=1.59; 1.18-2.14). Antibody detection was associated with residential exposures including living in crowded housing (RR=1.23; 0.98-1.53), with children (RR=1.40; 1.1-1.76) or unrelated roommates (RR=1.40; 1.19-1.64), and with a known or suspected COVID-19 case (RR=1.59; 1.13-2.24). Those who were obese (RR=1.65; 1.01-2.70) or diabetic (RR=1.31; 0.98-1.75) were also more likely to be seropositive. Farmworkers who lived in rural areas other than Greenfield (RR=0.58; 0.47-0.71), worked indoors (RR=0.68; 0.61-0.77), or whose employer provided them with information on how to protect themselves at work (RR=0.59; 0.40-0.86) had lower risk of prior infection.

**Conclusions and Relevance:** Our findings suggest both residential and workplace exposures are contributing to SARS-CoV-2 infection among farmworkers in California. Urgent distribution of COVID-19 vaccines is warranted given this population’s increased risk of infection and the essential nature of their work.

## INTRODUCTION

Essential workers in agriculture and food production have been severely affected by the ongoing COVID-19 pandemic. Widely-reported outbreaks associated with occupational exposure to SARS-CoV-2 have strained U.S. food supply chains^1^ and drawn attention to circumstances potentially placing workers at risk, including poor hygienic conditions, lack of access to educational materials in languages other than English, medical leave policies, and residential crowding.^2^

Agricultural work is one of the lowest-paid occupations of the U.S. economy, with 29% of full-time workers earning an annual income below $26,200 for a family of four.^3^ More than half of U.S. farmworkers are Latino,^4^ and many live in crowded housing of substandard quality.^5^ In California, at least half of farmworkers are believed to be undocumented, which could further lead to labor exploitation and a less protected working environment.^6^ In a study in Monterey County, California, we have reported 4-fold higher SARS-CoV-2 test positivity among farmworkers than the county population at large, and 24% higher test positivity among farmworkers than other individuals residing in the same communities.^7^ Statewide, agricultural and food workers experienced a 39% higher risk of all-cause death from March-October 2020 than during the same period in 2019 — a greater increase than any other occupational group;^8^ for workers of Latino backgrounds, the increase in all-cause mortality was 60%.^9^

Because specific exposures accounting for the high risk of infection among farmworkers remain poorly understood, there is uncertainty about what strategies can reduce risk of infection in this population.^10^ Here, we assessed sociodemographic, household, community, and workplace factors associated with SARS-CoV-2 infection and antibody response in a population of over 1,000 farmworkers working in the Salinas Valley in Monterey County, California.

## METHODS

### Study setting

The Salinas Valley, located within Monterey County, California, is home to an agricultural workforce of approximately 50,000 resident farmworkers, with an additional 40,000 seasonal workers supporting the peak summer and fall seasons.^11^ Clinica de Salud del Valle de Salinas (CSVS), a federally qualified community health center, is the main healthcare provider for Monterey County’s farmworkers and their families with a network of 12 clinics throughout the valley serving approximately 50,000 low-income, primarily Spanish-speaking patients.

### SARS-CoV-2 testing

Testing for SARS-CoV-2 infection at CSVS began 15 June 2020 and was offered to all individuals regardless of exposure, symptoms, documentation, or health insurance status. Medical personnel collected oropharyngeal specimens for detection of SARS-CoV-2 RNA via the qualitative Hologic (Marlborough, Massachusetts) Aptima nucleic acid transcription-mediated amplification (TMA) assay. Testing was conducted on clinic premises or at community sites, including low-income housing, agricultural fields, and CSVS-run community health fairs.

### Study enrollment

Between 16 July and 30 November 2020, we invited farmworkers (i.e., anyone employed in the agricultural sector) receiving care or getting tested for SARS-CoV-2 infection at CSVS clinics and community sites to participate in our study. We posted flyers about the study at the clinics and around town and provided study information to community groups and growers. Farmworkers were eligible for participation if they were not pregnant, ≥18 years old, had conducted farm work within the two weeks preceding their testing date, and were sufficiently proficient in English or Spanish to give consent and complete study procedures. Beginning 5 October, we enrolled any individuals who had engaged in agricultural work at any time since March 2020 since the growing season was ending.

We enrolled a total of 1,115 farmworkers. We excluded 8 farmworkers who did not provide blood samples or were not employed as farmworkers at time of enrollment from analyses, leaving a total of 1,107 participants. Study protocols were approved by the Office for the Protection of Human Subjects at UC Berkeley. All participants provided informed written consent.

### Study procedures

After the participant completed the SARS-CoV-2 TMA test and consented to participate in the study, the study team obtained a blood sample by venipuncture for testing of anti-SARS-CoV-2 antibody status. We then measured (with shoes on) height and weight using a digital scale. The study team administered a 45-minute computer-guided questionnaire by telephone in Spanish or English within two days before (for pre-consented participants) or after the enrollment visit, but before SARS-CoV-2 testing results were available. The questionnaire gathered information on socio-demographic characteristics, risk factors for SARS-CoV-2 infection, and impacts of the pandemic on daily life and wellbeing. Participants received a $50 prepaid VISA gift card which was loaded upon completion of all data collection activities.

Blood specimens were stored immediately at 4-7°C and centrifuged and aliquoted within 48 hours following collection. We used an in-house enzyme-linked immunosorbent assay (ELISA) to measure immunoglobulin G (IgG) reactivity to the SARS-CoV-2 spike and receptor binding domain proteins, as described previously.^7^ Prior validation of the assay used convalescent sera from hospitalized, mildly symptomatic, and asymptomatic PCR-positive cases as well as pre-2020 control specimens.

### Statistical analyses

We analyzed current (TMA-positivity) and historical (IgG antibody reactivity) SARS-CoV-2 infection separately. Analyses examining risk factors for TMA positivity included participants who worked in agriculture in the two weeks preceding enrollment (n=911); analyses for seropositivity included all farmworkers who provided a blood sample (n=1,058).

We performed bivariate analyses for a wide range of socio-demographic, household, community, and work-related characteristics known or suspected to be associated with SARS-CoV-2 infection (**Tables 1-4** and **eTables 1-4**) and assessed correlations between these characteristics (**eFigure 1**). We included covariates in multivariate models if there were >5 TMA positive or seropositive cases in each category, respectively, and a chi-square or t-test p-value<0.2 in bivariate analyses. Categorical risk factors were modeled as shown in **Tables 1-4**, with the exception of language spoken at home (modeled as indigenous language spoken at home — yes/no) and working in the fields (yes/no). Age, years in the U.S., and household size were modeled as continuous variables. We did not consider specific agricultural crops in multivariate analyses because farmworkers reported working in a variety of them. We used backward stepwise elimination (with a threshold of p<0.1) to select covariates for inclusion in final models.

**Table 1.** Socio-demographic and health-related risk factors for TMA positivity and seropositivity among farmworkers, Monterey County, 2020.

| Attribute | Current SARS-CoV-2 infection (TMA)<br>N=911 |  |  | Historical SARS-CoV-2 infection (IgG reactivity)<br>N=1058 |  |
| --- | --- | --- | --- | --- | --- |
|  | All enrolled<br>N=1107 | Positive<br>N=118 | Negative<br>N=793 | Positive<br>N=201 | Negative<br>N=857 |
|  | n (%) or<br>M ± SD | n (%) or<br>M ± SD | n (%) or<br>M ± SD | n (%) or<br>M ± SD | n (%) or<br>M ± SD |
| Recruitment site |  |  |  |  |  |
| Clinics | 561 (50.7) | 95 (18.5)* | 420 (81.6) | 97 (18.4) | 429 (81.6) |
| Community outreach | 546 (49.3) | 23 (5.8) | 373 (94.2) | 104 (19.6) | 428 (80.5) |
| Agricultural work in the preceding two weeks |  |  |  |  |  |
| No | 193 (17.4) | 23 (11.5) | 177 (88.5) | 156 (18.0)<sup>†</sup> | 148 (82.0) |
| Yes | 914 (82.6) | 118 (13.0) | 793 (87.1) | 45 (23.3) | 709 (76.7) |
| Sex |  |  |  |  |  |
| Female | 581 (52.5) | 60 (13.0) | 400 (87.0) | 99 (18.1) | 448 (81.9) |
| Male | 526 (47.5) | 58 (12.9) | 393 (87.1) | 102 (20.0) | 409 (80.0) |
| Age (years) | 39.7 ± 12.6 | 39.6 ± 11.0 | 39.6 ± 12.4 | 39.6 ± 12.3 | 39.6 ± 12.6 |
| 18-29 | 275 (24.8) | 27 (12.0) | 198 (88.0) | 43 (16.4)<sup>#</sup> | 220 (83.7) |
| 30-39 | 271 (24.5) | 29 (13.0) | 195 (87.1) | 59 (22.5) | 203 (77.5) |
| 40-49 | 297 (26.8) | 42 (16.4) | 214 (83.6) | 59 (20.8) | 225 (79.2) |
| 50-59 | 198 (17.9) | 16 (10.2) | 141 (89.8) | 27 (14.5) | 159 (85.5) |
| ≥60 | 66 (6.0) | 4 (8.2) | 45 (91.8) | 13 (20.6) | 50 (79.4) |
| Education |  |  |  |  |  |
| Primary school complete or less | 488 (44.1) | 63 (15.4)* | 346 (84.6) | 84 (21.3)* | 311 (78.7) |
| More than primary school | 618 (55.8) | 55 (11.0) | 446 (89.0) | 72 (15.4) | 397 (84.7) |
| No answer | 1 (0.1) | 0 (0.0) | 1 (100.0) | 0 (0.0) | 1 (100.0) |
| Marital status |  |  |  |  |  |
| Not married or living as married | 697 (63.0) | 50 (15.3)<sup>#</sup> | 277 (84.7) | 69 (17.8) | 319 (82.2) |
| Married or living as married | 409 (37.0) | 67 (11.5) | 516 (88.5) | 132 (19.7) | 537 (80.3) |
| No answer | 1 (0.1) | 1 (100.0) | 0 (0.0) | 0 (0.0) | 1 (100.0) |
| Annual household income |  |  |  |  |  |
| <\$25,000 | 557 (50.3) | 66 (14.5) | 390 (85.5) | 101 (18.8) | 435 (81.2) |
| ≥\$25,000 | 494 (44.6) | 48 (11.6) | 367 (88.4) | 86 (18.5) | 380 (81.6) |
| No answer | 56 (5.1) | 4 (10.0) | 36 (90.0) | 14 (25.0) | 42 (75.0) |
| Language spoken at home |  |  |  |  |  |
| Spanish | 940 (84.9) | 96 (12.4)* | 676 (87.6) | 166 (18.5) | 730 (81.5) |
| English | 57 (5.2) | 0 (0.0) | 42 (100.0) | 12 (21.8) | 43 (78.2) |
| Indigenous | 110 (9.9) | 22 (22.7) | 75 (77.3) | 23 (21.5) | 84 (78.5) |
| No answer | 0 (0.0) | 0 (0.0) | 0 (0.0) | 0 (0.0) | 0 (0.0) |
| Country of birth |  |  |  |  |  |
| Mexico | 922 (83.3) | 104 (13.5)<sup>#</sup> | 669 (86.5) | 163 (18.4) | 721 (81.6) |
| US | 141 (12.7) | 7 (7.0) | 93 (93.0) | 31 (23.0) | 104 (77.0) |
| Other | 44 (4.0) | 7 (18.4) | 31 (81.6) | 7 (18.0) | 32 (82.1) |
| Years in US | 21.3 ± 11.1 | 20.2 ± 10.9<sup>#</sup> | 21.0 ± 11.2 | 21.3 ± 10.6 | 21.2 ± 11.3 |
| <15 | 262 (23.7) | 38 (16.5)<sup>#</sup> | 193 (83.6) | 46 (18.0)<sup>#</sup> | 209 (82.0) |
| 15-19 | 191 (17.3) | 17 (10.4) | 147 (89.6) | 44 (24.0) | 139 (76.0) |
| 20-29 | 296 (26.7) | 34 (13.5) | 218 (86.5) | 49 (17.4) | 232 (82.6) |
| ≥30 | 216 (19.5) | 22 (13.4) | 142 (86.6) | 31 (15.3) | 172 (84.7) |
| Entire life | 141 (12.7) | 7 (7.0) | 93 (93.0) | 31 (23.0) | 104 (77.0) |
| No answer | 1 (0.1) | 0 (0.0) | 0 (0.0) | 0 (0.0) | 1 (100.0) |
| Community of residence |  |  |  |  |  |
| Salinas | 486 (43.9) | 40 (10.4)* | 343 (89.6) | 99 (21.2)* | 369 (78.9) |
| Greenfield | 315 (28.5) | 56 (19.8) | 227 (80.2) | 63 (21.2) | 234 (78.8) |
| Other town | 306 (27.6) | 22 (9.0) | 223 (91.0) | 39 (13.3) | 254 (86.7) |
| Smoking |  |  |  |  |  |
| Never smoked | 899 (81.2) | 99 (13.6) | 630 (86.4) | 157 (18.4) | 698 (81.6) |
| Former smoker | 158 (14.3) | 16 (11.4) | 124 (88.6) | 36 (23.4) | 118 (76.6) |
| Current smoker | 49 (4.4) | 3 (7.3) | 38 (92.7) | 8 (16.7) | 40 (83.3) |
| No answer | 1 (0.1) | 0 (0.0) | 1 (100.0) | 0 (0.0) | 1 (100.0) |
| Body mass index (measured) | 29.7 ± 5.5 | 29.2 ± 4.7 | 29.7 ± 5.6 | 30.4 ± 5.4* | 29.4 ± 5.5 |
| <25 (underweight or normal) | 197 (17.8) | 18 (10.8) | 149 (89.2) | 24 (12.5)* | 168 (87.5) |
| 25-29.9 (overweight) | 421 (38.0) | 45 (13.0) | 302 (87.0) | 75 (18.6) | 329 (81.4) |
| ≥30 (obese) | 461 (41.6) | 49 (13.1) | 326 (86.9) | 95 (21.7) | 342 (78.3) |
| Not collected | 28 (2.5) | 6 (27.3) | 16 (72.7) | 7 (28.0) | 18 (72.0) |
| Self-reported hypertension |  |  |  |  |  |
| No | 954 (86.2) | 106 (13.5) | 680 (86.5) | 171 (18.7) | 744 (81.3) |
| Yes | 149 (13.5) | 12 (9.8) | 110 (90.2) | 29 (20.9) | 110 (79.1) |
| No answer | 4 (0.4) | 0 (0.0) | 3 (100.0) | 1 (25.0) | 3 (75.0) |
| Self-reported diabetes |  |  |  |  |  |
| No | 977 (88.3) | 109 (13.6)<sup>#</sup> | 694 (86.4) | 172 (18.4)<sup>#</sup> | 762 (81.6) |
| Yes | 126 (11.4) | 9 (8.6) | 96 (91.4) | 28 (23.3) | 92 (76.7) |
| No answer | 4 (0.4) | 0 (0.0) | 3 (100.0) | 1 (25.0) | 3 (75.0) |

<sup>#</sup>p<0.2, <sup>†</sup>p<0.1, \*p<0.05

We used multiple imputation with chained equations to account for missing values (<2.5% missing for all variables). To account for differences between those recruited at clinics vs. community events, as well as changes in the background positivity rate in Monterey County over the course of the study period, we grouped participants into strata by recruitment site and period (i.e., 16 July-31 August, 1-30 September, 1-31 October, or 1-30 November). We used conditional fixed-effects Poisson models^12^ to estimate adjusted risk ratios (aRRs) while accounting for differences among strata, estimating robust standard errors using the Huber-White estimator.

## RESULTS

Most study participants were born in Mexico, had primary school or lower levels of educational attainment, were married or living as married, and were overweight or obese (**Table 1**). Participants averaged 39.7 (standard deviation (SD)=12.6) years of age and had lived in the U.S. for an average of 21.3 (SD=11.1) years. About 85% spoke Spanish at home, and 10% spoke an indigenous language. Half of participants reported household earnings <$25,000 per year. Three-quarters lived with children, including 36.9% living with children five years old or younger (**Table 2**). As many as 18.6% lived with unrelated roommates and 74.4% lived with other farmworkers. On average, farmworkers lived with 5.5 (SD=2.5) household members and 36.5% lived in crowded conditions (>2 persons/bedroom). Few reported leaving home for non-essential reasons (13%) or attending social gatherings with non-household members (10.1%) in the two weeks preceding the survey. As many as 17.9% reported living with someone who had symptoms of COVID-19 or were known to be infected with SARS-CoV-2 since the pandemic started, and 10.9% reported such exposures at home in the two weeks preceding their test.

**Table 2.** Household and community risk factors for TMA positivity and seropositivity among farmworkers, Monterey County, 2020.

| Attribute | Current SARS-CoV-2 infection (TMA)<br>N=911 |  |  | Historical SARS-CoV-2 infection (IgG reactivity)<br>N=1058 |  |
| --- | --- | --- | --- | --- | --- |
|  | All enrolled<br>N=1107 | Positive<br>N=118 | Negative<br>N=793 | Positive<br>N=201 | Negative<br>N=857 |
|  | n (%) or<br>M ± SD | n (%) or<br>M ± SD | n (%) or<br>M ± SD | n (%) or<br>M ± SD | n (%) or<br>M ± SD |
| Type of housing |  |  |  |  |  |
| House | 522 (47.2) | 60 (13.5) | 383 (86.5) | 101 (20.6) | 389 (79.4) |
| Apartment | 481 (43.6) | 48 (12.9) | 324 (87.1) | 85 (18.2) | 383 (81.8) |
| Hotel or motel | 37 (3.3) | 6 (16.7) | 30 (83.3) | 5 (13.5) | 32 (86.5) |
| Trailer or mobile home | 43 (3.9) | 3 (7.7) | 36 (92.3) | 8 (19.5) | 33 (80.5) |
| Other | 24 (2.2) | 1 (4.8) | 20 (95.2) | 2 (9.1) | 20 (90.9) |
| Household size | 5.5 ± 2.5 | 5.5 ± 2.4 | 5.4 ± 2.3 | 5.9 ± 2.6* | 5.4 ± 2.6 |
| 0 others | 12 (1.1) | 2 (18.2) | 9 (81.8) | 3 (25.0)* | 9 (75.0) |
| 1-3 others | 397 (35.9) | 41 (12.4) | 290 (87.6) | 58 (15.3) | 321 (84.7) |
| 4-6 others | 512 (46.3) | 51 (12.3) | 363 (87.7) | 93 (19.1) | 393 (80.9) |
| ≥7 others | 186 (16.8) | 24 (15.5) | 131 (84.5) | 47 (26.0) | 134 (74.0) |
| Children <18 living in the home | 1.8 ± 1.6 | 1.8 ± 1.5 | 1.8 ± 1.6 | 2.1 ± 1.7* | 1.7 ± 1.5 |
| No | 277 (25.0) | 28 (11.9) | 207 (88.1) | 43 (16.0)<sup>#</sup> | 225 (84.0) |
| Yes | 829 (74.9) | 90 (13.3) | 585 (86.7) | 157 (19.9) | 632 (80.1) |
| No answer | 1 (0.1) | 0 (0.0) | 1 (100.0) | 1 (100.0) | 0 (0.0) |
| Children ≤5 living in the home |  |  |  |  |  |
| No | 699 (63.1) | 79 (13.7) | 498 (86.3) | 110 (16.4)* | 559 (83.6) |
| Yes | 408 (36.9) | 39 (11.7) | 295 (88.3) | 91 (23.4) | 298 (76.6) |
| Children attending school/daycare |  |  |  |  |  |
| No | 1018 (92.0) | 105 (12.6) | 726 (87.4) | 184 (18.9) | 792 (81.2) |
| Yes | 85 (7.7) | 12 (15.8) | 64 (84.2) | 16 (20.5) | 62 (79.5) |
| No answer | 4 (0.4) | 1 (25.0) | 3 (75.0) | 1 (25.0) | 3 (75.0) |
| Living with unrelated roommates |  |  |  |  |  |
| No | 901 (81.4) | 93 (12.7) | 639 (87.3) | 156 (18.1)<sup>#</sup> | 704 (81.9) |
| Yes | 206 (18.6) | 25 (14.0) | 154 (86.0) | 45 (22.7) | 153 (77.3) |
| Living with other farmworkers |  |  |  |  |  |
| No | 281 (25.4) | 26 (11.6) | 198 (88.4) | 49 (18.4) | 218 (81.7) |
| Yes | 823 (74.4) | 92 (13.5) | 592 (86.6) | 151 (19.2) | 637 (80.8) |
| No answer | 3 (0.3) | 0 (0.0) | 3 (100.0) | 1 (33.3) | 2 (66.7) |
| Household crowding |  |  |  |  |  |
| ≤2 persons per bedroom | 703 (63.5) | 71 (12.3) | 505 (87.7) | 113 (17.0)* | 553 (83.0) |
| >2 persons per bedroom | 404 (36.5) | 47 (14.0) | 288 (86.0) | 88 (22.5) | 304 (77.6) |
| Access to washing machine at home |  |  |  |  |  |
| No | 411 (37.1) | 49 (14.2) | 296 (85.8) | 77 (19.5) | 318 (80.5) |
| Yes | 696 (62.9) | 69 (12.2) | 497 (87.8) | 124 (18.7) | 539 (81.3) |
| Left home for non-essential reasons (past 2 weeks) |  |  |  |  |  |
| No | 957 (86.5) | 104 (13.1) | 693 (87.0) | 169 (18.5) | 745 (81.5) |
| Yes | 144 (13.0) | 12 (11.0) | 97 (89.0) | 29 (21.0) | 109 (79.0) |
| No answer | 6 (0.5) | 2 (40.0) | 3 (60.0) | 3 (50.0) | 3 (50.0) |
| Used public transportation/ride share services (past 2 weeks) |  |  |  |  |  |
| No | 1039 (93.9) | 113 (13.2)<sup>#</sup> | 745 (86.8) | 189 (19.1) | 801 (80.9) |
| Yes | 62 (5.6) | 3 (6.3) | 45 (93.8) | 8 (14.5) | 53 (85.5) |
| No answer | 6 (0.5) | 2 (40.0) | 3 (60.0) | 3 (50.0) | 3 (50.0) |
| Attended social gatherings with non-household members (past 2 weeks) |  |  |  |  |  |
| No | 993 (89.7) | 105 (12.8) | 714 (87.2) | 181 (19.1) | 768 (80.9) |
| Yes | 112 (10.1) | 13 (14.4) | 77 (85.6) | 20 (18.5) | 88 (81.5) |
| No answer | 2 (0.2) | 0 (0.0) | 2 (100.0) | 0 (0.0) | 1 (100.0) |
| Attended indoor gatherings with non-household members (past 2 weeks) |  |  |  |  |  |
| No | 1046 (94.5) | 109 (12.7)<sup>#</sup> | 753 (87.4) | 192 (19.2) | 807 (80.8) |
| Yes | 59 (5.3) | 9 (19.2) | 38 (80.9) | 9 (15.5) | 49 (84.5) |
| No answer | 2 (0.2) | 0 (0.0) | 2 (100.0) | 0 (0.0) | 1 (100.0) |
| Face covering use while less than 6 feet away from others all of the time |  |  |  |  |  |
| No | 85 (7.7) | 4 (10.5) | 34 (89.5) | 13 (16.1) | 68 (84.0) |
| Yes | 1022 (92.3) | 9 (17.3) | 43 (82.6) | 188 (19.2) | 789 (80.8) |
| Hand washing when returning home or after touching something all or most of the time |  |  |  |  |  |
| No | 33 (3.0) | 3 (11.1) | 24 (88.9) | 5 (16.1) | 26 (83.9) |
| Yes | 1074 (97.0) | 115 (13.0) | 769 (87.0) | 196 (19.1) | 831 (80.9) |
| Possible exposure to someone with COVID-19 at home in the past 2 weeks<sup>a</sup> |  |  |  |  |  |
| No | 986 (89.1) | 79 (9.8)* | 725 (90.2) | 178 (18.8) | 767 (81.2) |
| Yes | 121 (10.9) | 39 (36.5) | 68 (63.6) | 23 (20.4) | 90 (70.7) |
| Possible exposure to someone with COVID-19 at home since the start of the pandemic<sup>b</sup> |  |  |  |  |  |
| No | 909 (82.1) | 68 (9.2)* | 675 (90.9) | 152 (17.4)* | 723 (82.6) |
| Yes | 198 (17.9) | 50 (29.8) | 118 (70.2) | 49 (26.8) | 134 (73.2) |

<sup>#</sup>p<0.2, <sup>†</sup>p<0.1, \*p<0.05
<sup>a</sup>Lived with someone who had COVID-19 symptoms or positive in the past 2 weeks.
<sup>b</sup>Lived with someone who had COVID-19 symptoms or positive since the start of the pandemic.

About three-quarters of participants worked in the fields and farmed a variety of crops; the most common were berries (28.6%), leafy greens (26.4%), and broccoli (18.8%) (**Table 3**). Nearly all farmworkers reported using a face covering at work, and 34.3% commuted to work with members of other households. Almost 40% worked during the pandemic with someone who had symptoms of COVID-19 or were known to be infected with SARS-CoV-2 and 13.5% reported such workplace exposure during the two weeks preceding their testing date. Almost all farmworkers reported that their employers provided them with hand sanitizer, gloves, face coverings, and handwashing stations; disinfected surfaces and tools regularly; and provided them with information on how to prevent SARS-CoV-2 transmission at work and the importance of staying away from work if they were sick (**Table 4**). However, 44.7% reported that their employer did not screen for fever and symptoms upon arrival at the workplace, which was recommended as part of a countywide agricultural advisory.^13^

**Table 3.** Work-related risk factors for TMA positivity and seropositivity among farmworkers, Monterey County, 2020.

| Attribute | Current SARS-CoV-2 infection (TMA) |  |  | Historical SARS-CoV-2 infection (IgG reactivity) |  |
| --- | --- | --- | --- | --- | --- |
|  |  | N=911 |  | N=1058 |  |
|  | All enrolled<br>N=1107 | Positive<br>N=118 | Negative<br>N=793 | Positive<br>N=201 | Negative<br>N=857 |
|  | n (%) or<br>M ± SD | n (%) or<br>M ± SD | n (%) or<br>M ± SD | n (%) or<br>M ± SD | n (%) or<br>M ± SD |
| H2A visa holder |  |  |  |  |  |
| No | 1029 (93.0) | 107 (12.7) | 733 (87.3) | 188 (19.2) | 792 (80.8) |
| Yes | 65 (5.9) | 9 (15.0) | 51 (85.0) | 11 (16.9) | 54 (83.1) |
| No answer | 13 (1.2) | 2 (18.2) | 9 (81.8) | 2 (15.4) | 11 (84.6) |
| Supervisor or mayordomo |  |  |  |  |  |
| No | 1015 (76.2) | 111 (12.8) | 756 (87.2) | 183 (18.9) | 784 (81.1) |
| Yes | 50 (4.5) | 7 (15.9) | 37 (84.1) | 9 (18.4) | 40 (81.6) |
| No answer | 42 (3.8) | 0 (0.0) | 0 (0.0) | 9 (21.4) | 33 (78.6) |
| Type of agricultural work<br>(ever/in the past 2 weeks)<sup>a</sup> |  |  |  |  |  |
| Worked in the fields | 825 (74.5) | 100 (14.7)* | 580 (85.3) | 162 (20.4)* | 633 (79.6) |
| Packing shed | 133 (12.0) | 11 (10.5) | 94 (89.5) | 21 (16.4) | 107 (83.6) |
| Processing facility | 63 (5.7) | 4 (7.0)<sup>#</sup> | 53 (93.0) | 7 (12.1)<sup>#</sup> | 51 (87.9) |
| Nursery | 38 (3.4) | 4 (12.1) | 29 (87.9) | 4 (11.4) | 31 (88.6) |
| Truck driver | 38 (3.4) | 4 (12.1) | 29 (87.9) | 3 (9.1)<sup>#</sup> | 30 (90.9) |
| Packing truck | 22 (2.0) | 1 (4.8) | 20 (95.2) | 2 (9.5) | 19 (90.5) |
| Other | 21 (1.9) | 1 (5.3) | 18 (94.7) | 2 (10.5) | 17 (89.5) |
| No answer | 10 (0.9) | 0 (0.0) | 1 (100.0) | 2 (20.0) | 8 (80.0) |
| Worked indoors |  |  |  |  |  |
| No | 844 (76.2) | 98 (14.3)* | 589 (85.7) | 166 (20.4)* | 646 (79.6) |
| Yes | 262 (23.7) | 20 (9.0) | 203 (91.0) | 35 (14.3) | 210 (85.7) |
| No answer | 1 (0.1) | 0 (0.0) | 1 (100.0) | 0 (0.0) | 1 (100.0) |
| Crops (ever/in the past 2<br>weeks)<sup>a</sup> |  |  |  |  |  |
| Berries | 236 (28.6) | 14 (7.2)* | 181 (92.8) | 39 (16.7)<sup>#</sup> | 194 (83.3) |
| Leafy greens | 218 (26.4) | 31 (17.9)<sup>#</sup> | 142 (82.1) | 50 (24.2)<sup>#</sup> | 157 (75.9) |
| Broccoli | 155 (18.8) | 28 (18.9)<sup>#</sup> | 120 (81.1) | 24 (16.3)<sup>#</sup> | 123 (83.7) |
| Grapes | 59 (7.2) | 10 (21.3)<sup>#</sup> | 37 (78.7) | 15 (26.3) | 42 (73.7) |
| Peas | 52 (6.3) | 16 (30.8)* | 36 (69.2) | 5 (10.0)<sup>†</sup> | 45 (90.0) |
| Cauliflower | 42 (5.1) | 5 (13.5) | 32 (86.5) | 11 (30.6)<sup>#</sup> | 25 (69.4) |
| Celery | 19 (2.3) | 2 (11.8) | 15 (88.2) | 3 (18.8) | 13 (81.3) |
| Artichokes | 6 (0.7) | 0 (0.0) | 5 (100.0) | 1 (16.7) | 5 (83.3) |
| Other | 158 (19.2) | 9 (8.4)* | 98 (91.6) | 35 (22.6) | 120 (77.4) |
| Commuted to work with non-household members |  |  |  |  |  |
| No | 707 (63.9) | 65 (10.9)* | 530 (89.1) | 124 (18.4) | 548 (81.6) |
| Yes | 380 (34.3) | 53 (16.8) | 263 (83.2) | 72 (19.7) | 294 (80.3) |
| No answer | 20 (1.8) | 0 (0.0) | 0 (0.0) | 5 (25.0) | 15 (75.0) |
| Used face covering at work all<br>of the time |  |  |  |  |  |
| No | 112 (10.1) | 11 (11.7) | 83 (88.3) | 19 (18.1) | 86 (81.9) |
| Yes | 992 (89.6) | 107 (13.1) | 709 (86.9) | 182 (19.2) | 768 (80.8) |
| No answer | 3 (0.3) | 0 (0.0) | 1 (100.0) | 0 (0.0) | 3 (100.0) |
| Came within 6 feet from others<br>while working |  |  |  |  |  |
| No | 501 (45.3) | 52 (12.6) | 361 (87.4) | 95 (19.7) | 388 (80.3) |
| Yes | 568 (51.3) | 64 (13.4) | 413 (86.6) | 97 (18.0) | 441 (82.0) |
| No answer | 38 (3.4) | 2 (9.5) | 19 (90.5) | 9 (24.3) | 28 (75.7) |
| Possible exposure to someone with COVID-19 at work in the past 2 weeks<sup>b</sup> |  |  |  |  |  |
| No | 958 (86.5) | 83 (10.9)* | 680 (89.1) | 178 (19.3) | 744 (80.7) |
| Yes | 149 (13.5) | 35 (23.7) | 113 (76.4) | 23 (16.9) | 113 (83.1) |
| No answer | 0 (0.0) | 0 (0.0) | 0 (0.0) | 0 (0.0) | 0 (0.0) |
| Possible exposure to someone with COVID-19 at work since the start of the pandemic<sup>c</sup> |  |  |  |  |  |
| No | 665 (60.1) | 52 (9.9)* | 471 (90.1) | 117 (18.3) | 524 (81.8) |
| Yes | 442 (39.9) | 66 (17.0) | 322 (83.0) | 84 (20.1) | 333 (79.9) |
| No answer | 0 (0.0) | 0 (0.0) | 0 (0.0) | 0 (0.0) | 0 (0.0) |

**Table 4.**
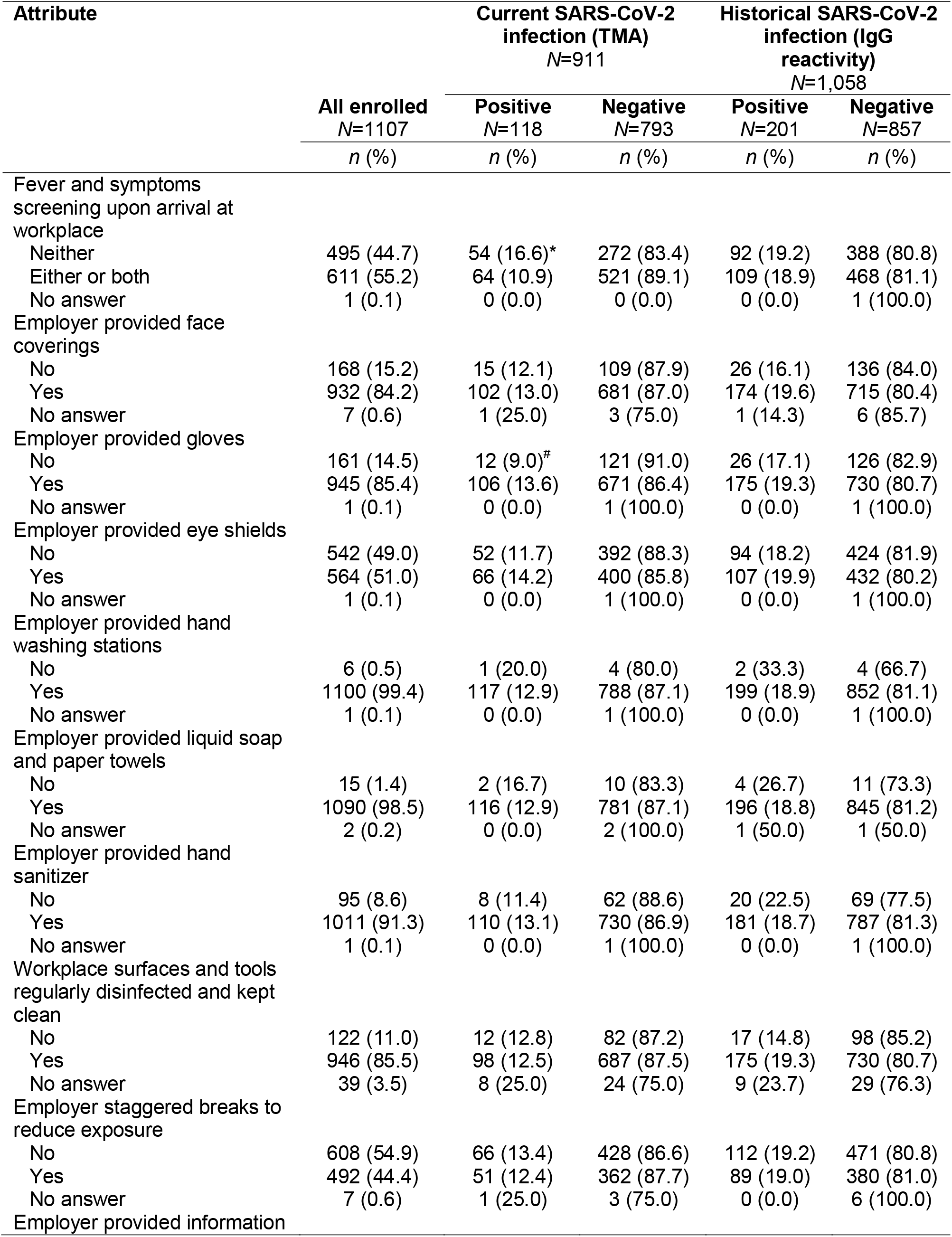
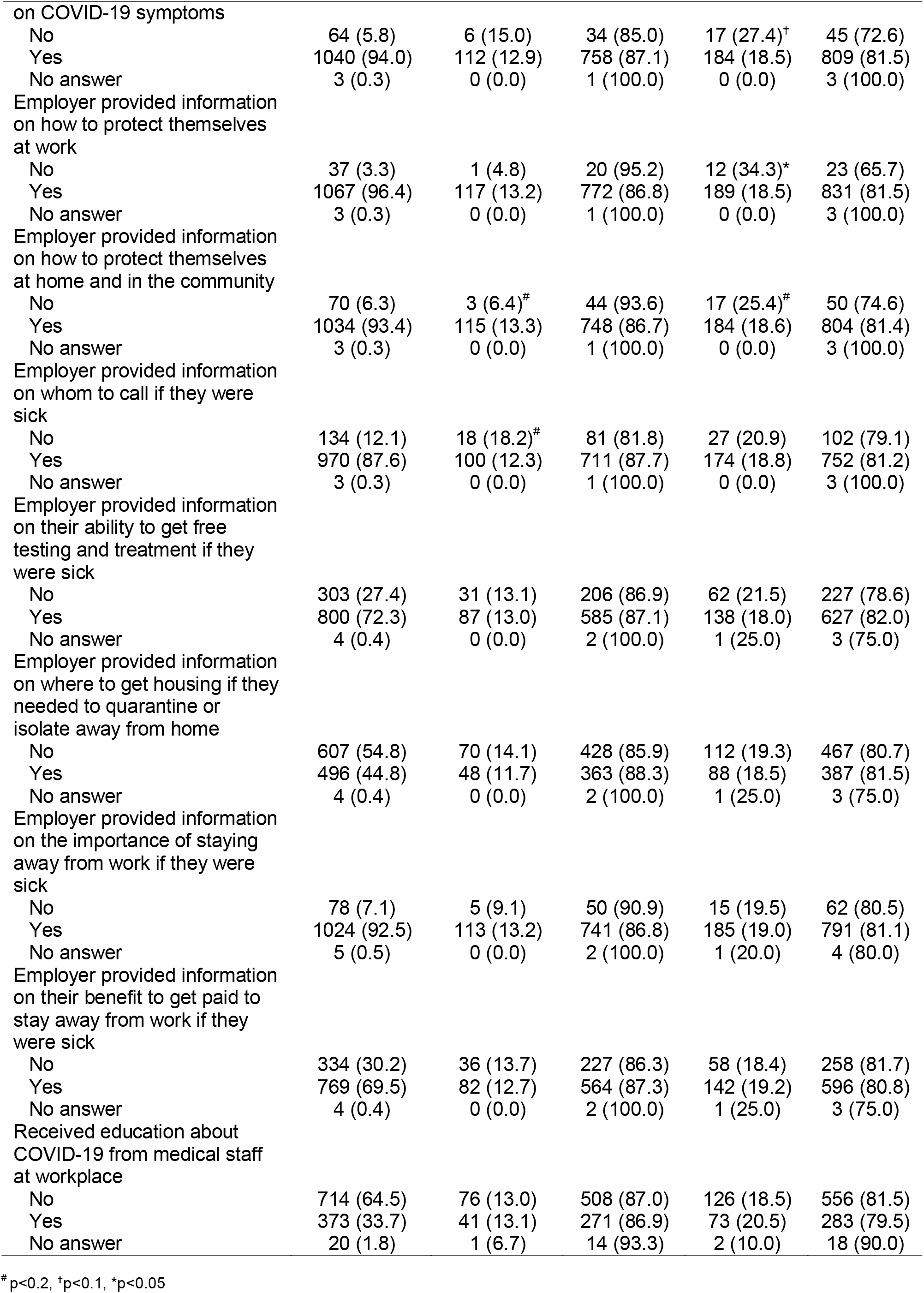
Employer-provided preventive measures and their association with TMA positivity and seropositivity among farmworkers, Monterey County, 2020.

### Risk factors for current SARS-CoV-2 infection (TMA-positivity)

Thirteen percent (n=118) of the 911 participants currently working in agriculture tested positive for current SARS-CoV-2 shedding by TMA, including 18.5% of those recruited at the clinics and 5.8% of those recruited via outreach (**Table 1**). Several socio-demographic, household, community, and work-related variables were associated with TMA positivity in bivariate analyses (**Tables 1-4**). Notably, we found that participants who had a lower educational level, spoke indigenous languages at home, lived in the community of Greenfield, worked in the fields, did not work indoors, commuted to work with non-household members, lived or worked with someone who had symptoms of COVID-19 or with known infection in the preceding two weeks, and were not screened for either fever or COVID-19 symptoms upon arrival at work had a higher prevalence of TMA positivity. We also observed correlations between some of these characteristics (**eFigure 1**).

In multivariate analyses, we found that TMA positivity was more common among individuals who had only primary school or no education (aRR=1.32; 95% CI: 0.99-1.76), spoke an indigenous language at home (aRR=1.30; 0.97-1.73), or lived with (aRR=2.98; 2.06-4.32) or worked with (aRR=1.59; 1.18-2.14) someone who had symptoms of COVID-19 or was known to be infected with SARS-CoV-2 in the previous two weeks. Additionally, working in the fields (compared to agricultural work in all other settings) was associated with higher risk of current SARS-CoV-2 infection (aRR=1.60; 1.03-2.50). In contrast, farmworkers screened by employers for symptoms of COVID-19 or elevated temperature had a reduced risk of current infection (aRR=0.79; 0.61-1.01; **Figure 1A**).

**Figure 1.**
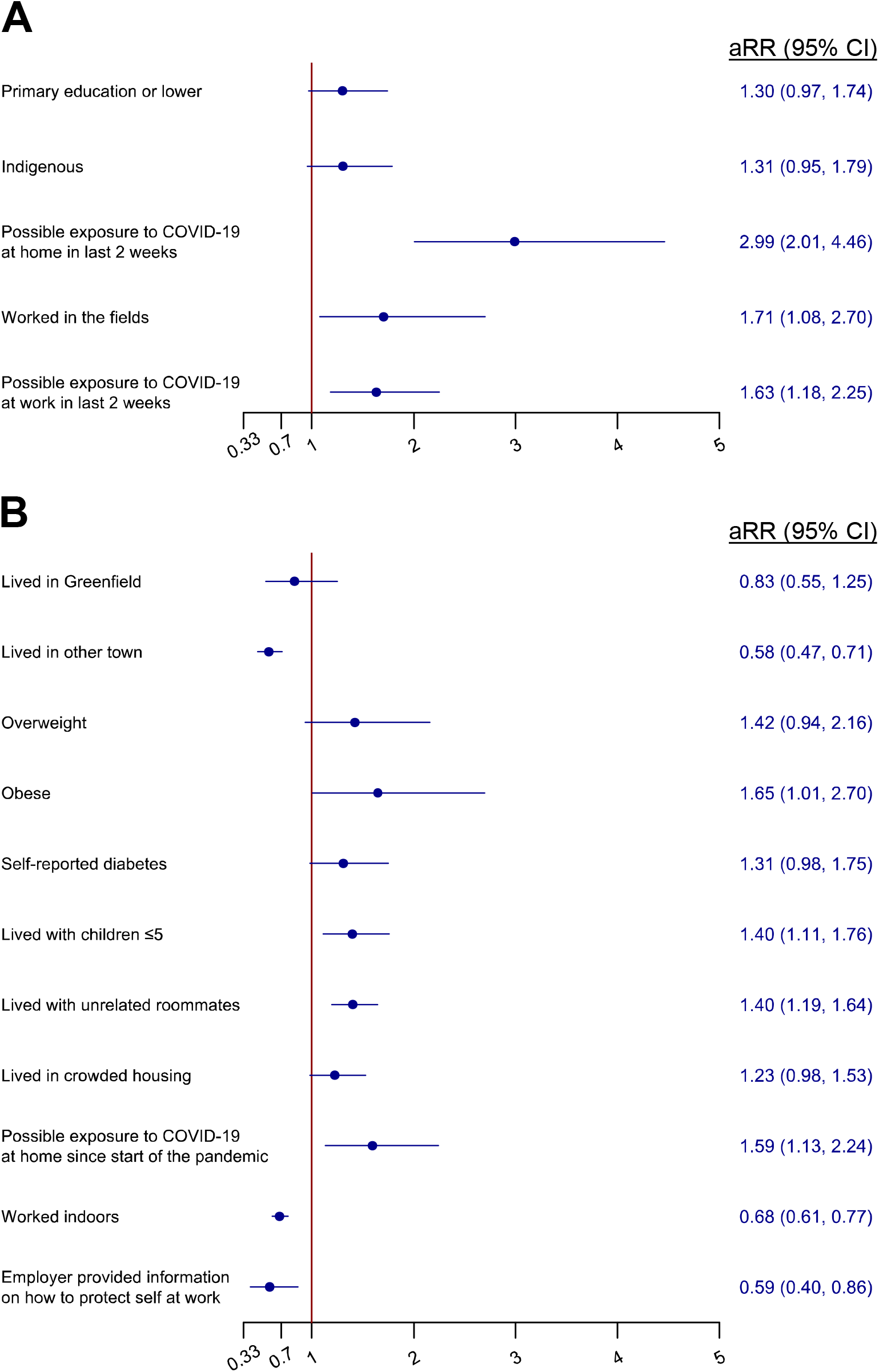
Change in: (**A**) TMA-positivity risk for explanatory variables among current farmworkers (n=911), and (**B**) seropositivity risk for explanatory variables among all farmworkers who provided a blood sample (n=1,058). Adjusted relative risks (aRR) and 95% confidence intervals from conditional fixed-effects Poisson models.

### Risk factors for prior SARS-CoV-2 infection (IgG antibody reactivity)

We found that 19% of the participants who provided a blood sample had antibody evidence of prior infection, with similar prevalence among those tested in the clinics (18.4%) and at community sites (19.4%; **Table 1**). Farmworkers with primary school or no education, who lived in Salinas or Greenfield (vs. other towns), were overweight or obese, lived in large households or with children ≤5 years, lived in crowded housing, had ever lived with someone who had symptoms of COVID-19 or were known to be infected with SARS-CoV-2, and worked in the fields had higher antibody prevalence than their counterparts (**Tables 1-3**). We also found that those who worked indoors and whose employer provided information on how to protect themselves at work had lower likelihood of seropositivity (**Tables 3** and **4**).

In multivariate analyses, we found that participants who were obese (aRR=1.65; 1.01-2.70) or overweight (aRR=1.42; 0.94-2.16) or reported being diabetic (aRR=1.31; 0.98-1.75) were more likely to be seropositive. Additionally, we identified higher risk of seropositive status among those living with children ≤ 5 years old (aRR=1.40; 1.1-1.76), with unrelated roommates (aRR=1.40; 1.19-1.64) or in crowded housing (aRR=1.23; 0.98-1.53), and those who had ever lived with someone who had symptoms of COVID-19 or were known to be infected with SARS-CoV-2 (aRR=1.59; 1.13-2.24; **Figure 1B**).

Farmworkers who lived outside the region’s largest communities of Salinas and Greenfield (aRR=0.58; 0.47-0.71), worked indoors (aRR=0.68; 0.61-0.77), or whose employer provided them with information on how to protect themselves at work (aRR=0.59; 0.40-0.86) had a decreased risk of serpositivity.

## DISCUSSION

In this primarily Mexican-born and very low-income farmworker population in California, current SARS-CoV-2 infection was associated with having lower levels of education, speaking an indigenous language, working in the fields rather than elsewhere in agriculture, and exposure to known or suspected COVID-19 case at home or in the workplace. We also found higher prevalence of prior SARS-CoV-2 infection, indicated by IgG seropositivity, associated with outdoor work and with residential exposures (living in crowded housing, with children or unrelated roommates, and with a known or suspected COVID-19 case). Those living in the more urban areas of the county were particularly at risk, as were those who were obese or diabetic. As evidence of the importance of health education, farmworkers who reported that their employer provided them with information on COVID-19 protection were less likely to have been infected.

Our study suggests several routes of SARS-CoV-2 exposure that may be of importance to the farmworker population. Unsurprisingly, living in crowded housing or with unrelated roommates was associated with higher risk of prior infection. Independent of these associations, we also saw higher seroprevalence among individuals living with children five years old or younger. While the role of children in SARS-CoV-2 transmission has been uncertain in many populations, in part due to lower risk of symptoms and lower frequency of testing at younger ages,^14–16^ recent investigations have demonstrated equivalent viral load across ages^17^ and higher risk of transmission from infected children than from adults, given similar household exposures.^18^ While schools and formal daycare establishments were closed during our study, informal or home-based childcare arrangements with relatives or friends may have led to additional exposure to infection. Taken together, our findings suggest substantial risk of infection associated with residential exposures in this low-income essential workforce population.

Several workplace factors were also associated with infection risk. Farmworkers whose employers provided informational resources on preventing COVID-19 at work had 41% lower risk of prior infection, whereas farmworkers whose employers screened them for symptoms or fever had 21% lower risk of current infection. This reduction could owe to benefits of health education, as well as more stringent efforts by employers to reduce risk by providing education and screenings. Individuals working outside and in the fields were more likely to be both currently infected and seropositive, respectively. Whereas indoor exposures are thought to be associated with the greatest risk of transmission,^19^ a lower perceived sense of risk during outdoor work, difficulty using PPE while engaged in physically demanding tasks, or socioeconomic differences among outdoor and indoor workers may contribute to the observed association in our study. While the estimated risk ratio for infection associated with workplace exposure was lower than that for household exposure, this difference could in part reflect misclassification, if individuals are are more likely to know about the health of household members. Previously, we have reported higher SARS-CoV-2 test positivity among farmworkers than among age-and sex-matched adults from the same communities who also received testing at CSVS,^7^ further supporting the hypothesis that workplace exposures specific to agriculture may be of importance to SARS-CoV-2 transmission.

Last, we observed that farmworkers who spoke indigenous languages and those with lower education were more likely to currently have COVID-19 at the time of testing. Those who spoke indigenous languages also had a lower educational level and had more recently arrived in the U.S. They lived in more crowded conditions and were more likely to work in the fields and to commute to work with non-household members (see **eFigure 1**). Only limited COVID-19 health messages have been provided in indigenous languages, which are primarily not written languages.

We also saw associations of prior infection with comorbid conditions. While it is known that obesity increases the risk for severe COVID-19 illness,^20^ we also observed an increased risk of prior infection among obese individuals. This finding is consistent with a recent meta-analysis of 20 studies which found 46% higher odds of SARS-CoV-2 infection among obese individuals,^20^ possibly related to alterations in systemic metabolism, including altered adipokines^21–23^ and chronic low-grade inflammation.^24,25^ Similarly, diabetes can attenuate the synthesis of proinflammatory cytokines (e.g., interferon gamma and interleukins) and their downstream acute phase reactants,^26^ but also impair macrophage and lymphocyte functions.^27^ As obesity and diabetes are prevalent among farmworkers as well as other low-income Latino populations, our findings that these conditions are associated with higher risk of infection add to previous concerns based on the knowledge that these conditions may also exacerbate risk of adverse clinical outcomes.

Our work represents the first epidemiological study to address risk factors for SARS-CoV-2 infection among U.S. farmworkers, and substantiates earlier concerns^6,28–30^ that living and working conditions in this population may contribute to risk of infection. Several limitations should be considered. We cannot determine how well our sample represents the farmworker population, many of whom are “hidden” due to their informal workforce participation and undocumented status.^31^ As we excluded individuals who did not speak Spanish or English sufficiently well to participate, our study likely under-represents indigenous populations. We observed differences in prevalence of current, but not prior, SARS-CoV-2 infection between study participants recruited at clinics and those recruited via community outreach events,^7^ as individuals seeking testing at clinics were more likely to be symptomatic or to report recent known exposure; to mitigate confounding, we defined strata by recruitment site. In addition, waning antibodies, particularly for individuals experiencing mild or asymptomatic infection,^32^ may have contributed to misclassification for individuals infected early in the pandemic. Lastly, many identified risk factors were highly correlated, making it difficult to separate out their unique effects.

Our previous analyses demonstrated a high prevalence of SARS-CoV-2 infection among farmworkers in California’s Salinas Valley; findings reported here further underscore the urgent need to intervene on modifiable risk factors such as increasing availability of isolation facilities to reduce exposure to COVID-19 cases at home and access to paid medical leave to avoid transmission in the workplace. Farmworkers speaking indigenous languages, who have very low levels of formal education, and who live in rural communities were at especially high risk of infection in our study, demonstrate disparities even within this very low-income and high-risk population. Efficacious vaccines have now been authorized and should be distributed to farmworkers with urgency owing to the high risk of infection in this population, and the essential nature of their work.

## Data Availability

Direct data requests to the corresponding author at.

## ACKNOWLEDGEMENTS

This work was supported by the Innovative Genomics Institute and Clinica de Salud del Valle de Salinas. JAL discloses receipt of grants and fees from Pfizer unrelated to this study. All other authors declare no conflicts of interest.

The study was reviewed and approved by the Committee for Protection of Human Subjects at UC Berkeley.

Members of the CHAMACOS-Project-19 Study Team include Jose Camacho, Gardenia Casillas, Celeste Castro, Madison J. de Vere, Lupe Flores, Lizari Garcia, Maria Reina Garcia, Terry Gomez, Carly Hyland, Daniel Lampert, Aaron McDowell-Sanchez, Dominic Pina Montes, Jacqueline Montoya, Oguchi Nkwocha, Lilibeth Nunez, Juanita “Liz” Orozco, Marbel Orozco, Kimberly L. Parra, Nargis Rezai, Maria T. Rodriquez, Monica Romero, Hina Sheth, Jon Yoshiyama, and Litzi Zepeda.

