## Supplemental Material for "Risk factors for SARS-CoV-2 infection among farmworkers in Monterey County, California"

Table of contents

| **Supplemental table/figure** | **Page number** |
| --- | --- |
| eTable 1. Sociodemographic and health-related risk factors for TMA positivity and seropositivity among farmworkers, Monterey County, 2020. | 2 |
| eTable 2. Household and community risk factors for TMA positivity and seropositivity among farmworkers, Monterey County, 2020. | 4 |
| eTable 3. Work-related risk factors for TMA positivity and seropositivity among farmworkers, Monterey County, 2020. | 7 |
| eTable 4. Employer-provided preventive measures and their association with TMA positivity and seropositivity among farmworkers, Monterey County, 2020. | 9 |
| eFigure 1. Correlation heat map of risk factors related to TMA positivity or seropositivity among farmworkers, Monterey County, 2020. | 12 |

| **eTable 1.** Sociodemographic and health-related risk factors for TMA positivity and seropositivity among farmworkers, Monterey County, 2020. | | | | | | | |
| --- | --- | --- | --- | --- | --- | --- | --- |
| **Attribute** | **Current SARS-CoV-2 infection (TMA)** | | |  | **Historical SARS-CoV-2 infection (IgG reactivity)** | | |
|  | All enrolled  N=911 | Positive  N=118 | Negative  N=793 |  | All enrolled  N=1058 | Positive  N=201 | Negative  N=857 |
|  | *n* (%) or M±SD | *n* (%) or M±SD | *n* (%) or M±SD |  | *n* (%) or M±SD | *n* (%) or M±SD | *n* (%) or M±SD |
| Recruitment site |  |  |  |  |  |  |  |
| Clinics | 515 (56.5) | 95 (80.5) | 420 (53.0) |  | 526 (49.7) | 97 (48.3) | 429 (50.1) |
| Community outreach | 396 (43.5) | 23 (19.5) | 373 (47.0) |  | 532 (50.3) | 104 (51.7) | 428 (49.9) |
| Agricultural work in the preceding two weeks |  |  |  |  |  |  |  |
| No | NA | NA | NA |  | 193 (18.2) | 156 (77.6) | 709 (82.7) |
| Yes | 911 (100.0) | 118 (100.0) | 793 (100.0) |  | 865 (81.8) | 45 (22.4) | 148 (17.3) |
| Sex |  |  |  |  |  |  |  |
| Female | 460 (50.5) | 60 (50.9) | 393 (49.6) |  | 547 (51.7) | 99 (49.3)) | 448 (52.3) |
| Male | 451 (49.5) | 58 (49.2) | 400 (50.4) |  | 511 (48.3) | 102 (50.8) | 409 (47.7) |
| Age (years) | 39.6 ± 12.2 | 39.6 ± 11.0 | 39.6 ± 12.4 |  | 39.6 ± 12.6 | 30.4 ± 5.4 | 39.6 ±12.6 |
| 18-29 | 225 (24.7) | 27 (22.9) | 198 (25.0) |  | 263 (24.9) | 43 (21.4) | 220 (25.7) |
| 30-39 | 224 (24.6) | 29 (24.6) | 195 (24.6) |  | 262 (24.8) | 59 (29.4) | 203 (24.0) |
| 40-49 | 256 (28.1) | 42 (35.6) | 214 (27.0) |  | 284 (26.8) | 59 (29.4) | 225 (26.3) |
| 50-59 | 157 (17.2) | 16 (13.6) | 141 (17.9) |  | 186 (17.6) | 27 (13.4) | 159 (18.6) |
| ≥60 | 49 (5.4) | 4 (3.4) | 45 (5.7) |  | 63 (6.0) | 13 (6.5) | 50 (5.8) |
| Education |  |  |  |  |  |  |  |
| Primary school complete or less | 409 (44.9) | 63 (46.6) | 346 (43.6) |  | 585 (55.3) | 100 (50.0) | 484 (56.5) |
| More than primary school | 501 (55.0) | 55 (53.6) | 446 (56.2) |  | 472 (44.6) | 101(50.3) | 372 (43.4) |
| No answer | 1 (0.1) | 0 (0.0) | 1 (0.1) |  | 1 (0.1) | 0 (0.0) | 1 (0.1) |
| Marital status |  |  |  |  |  |  |  |
| Not married or living as married | 327 (35.9) | 50 (42.4) | 277 (34.9) |  | 388 (36.7) | 69 (34.3) | 319 (37.2) |
| Married or living as married | 583 (64.0) | 67 (56.8) | 516 (65.1) |  | 669 (63.2) | 132 (65.7) | 537 (62.7) |
| No answer | 1 (0.1) | 1 (0.9) | 0 (0.0) |  | 1 (0.1) | 0 (0.0) | 1 (0.1) |
| Annual household income |  |  |  |  |  |  |  |
| <$25,000 | 456 (50.1) | 66 (55.9) | 390 (49.2) |  | 536 (50.7) | 101 (50.3) | 435 (50.8) |
| ≥$25,000 | 415 (45.6) | 48 (40.7) | 367 (46.3) |  | 466 (44.1) | 86 (42.8) | 380 (44.3) |
| No answer | 40 (4.4) | 4 (3.4) | 36 (4.5) |  | 56 (5.3) | 14 (7.0) | 42 (4.9) |
| Language spoken at home |  |  |  |  |  |  |  |
| Spanish | 772 (84.7) | 96 (81.4) | 676 (85.2) |  | 896 (84.7) | 166 (82.6) | 730 (85.2) |
| English | 42 (4.6) | 0 (0.0) | 42 (5.3) |  | 55 (5.2) | 12 (6.0) | 43 (5.0) |
| Indigenous | 97 (10.7) | 22 (18.6) | 75 (9.4) |  | 107 (10.1) | 23 (11.4) | 84 (9.8) |
| No answer | 0 (0.0) | 0 (0.0) | 0 (0.0) |  | 0 (0.0) | 0 (0.0) | 0 (0.0) |
| Country of birth |  |  |  |  |  |  |  |
| Mexico | 773 (84.9) | 104 (88.1) | 669 (84.3) |  | 884 (83.6) | 163 (81.1) | 721 (83.1) |
| US | 100 (11.0) | 7 (5.9) | 93 (11.7) |  | 135 (12.8) | 31 (15.4) | 104 (12.1) |
| Other | 38 (4.2) | 7 (5.9) | 31 (3.9) |  | 39 (3.7) | 7 (3.5) | 32 (3.7) |
| Years in US | 20.9 ± 11.1 |  |  |  | 21.3 ± 11.1 |  |  |
| <15 | 231 (25.4) | 38 (32.2) | 193 (24.3) |  | 255 (24.1) | 46 (22.9) | 209 (24.4) |
| 15-19 | 164 (18.0) | 17 (14.4) | 147 (18.5) |  | 183 (17.3) | 44 (21.9) | 139 (16.2) |
| 20-29 | 252 (27.7) | 34 (28.8) | 218 (27.5) |  | 281 (26.6) | 49 (24.4) | 232 (27.1) |
| ≥30 | 164 (18.0) | 22 (18.6) | 142 (17.9) |  | 203 (19.2) | 31 (15.4) | 172 (20.1) |
| Entire life | 100 (11.0) | 7 (5.9) | 93 (11.7) |  | 135 (12.8) | 31 (15.4) | 104 (12.1) |
| No answer | 0 (0.0) | 0 (0.0) | 0 (0.0) |  | 1 (0.1) | 0 (0.0) | 1 (0.1) |
| Community of residence |  |  |  |  |  |  |  |
| Salinas | 383 (42.0) | 40 (33.9) | 343 (43.3) |  | 468 (44.2) | 99 (49.3) | 369 (34.1) |
| Greenfield | 283 (31.1) | 56 (47.5) | 227 (28.6) |  | 297 (28.1) | 63 (31.3) | 234 (27.3) |
| Other town | 245 (26.9) | 22 (18.6) | 223 (28.1) |  | 293 (27.7) | 39 (19.4) | 254 (29.6) |
| Smoking |  |  |  |  |  |  |  |
| Never smoked | 729 (80.0) | 99 (83.9) | 630 (79.5) |  | 855 (80.8) | 157 (78.1) | 698 (81.5) |
| Former smoker | 140 (15.4) | 16 (13.6) | 124 (15.6) |  | 154 (14.6) | 36 (17.9) | 118 (13.8) |
| Current smoker | 41 (4.5) | 3 (2.5) | 38 (4.8) |  | 48 (4.5) | 8 (4.0) | 40 (4.7) |
| No answer | 1 (0.1) | 0 (0.0) | 1 (0.1) |  | 1 (0.1) | 0 (0.0) | 1 (0.1) |
| Body mass index (measured) | 29.6 ± 5.5 | 29.2 ± 4.7 | 29.7 ± 5.6 |  | 29.6 ± 5.5 | 30.4 ± 5.4 | 29.4 ± 5.5 |
| <25 (underweight or normal) | 167 (18.3) | 18 (15.3) | 149 (18.8) |  | 192 (18.2) | 24 (11.9) | 168 (19.6) |
| 25-29.9 (overweight) | 347 (38.1) | 45 (38.1) | 302 (38.0) |  | 404 (38.2) | 75 (37.3) | 329 (38.4) |
| ≥30 (obese) | 375 (41.2) | 49 (41.5) | 326 (41.1) |  | 437 (41.3) | 95 (47.3) | 342 (39.9) |
| Not collected | 22 (2.4) | 6 (5.1) | 16 (2.0) |  | 25 (2.4) | 7 (3.5) | 18 (2.1) |
| Self-reported hypertension |  |  |  |  |  |  |  |
| No | 786 (86.3) | 106 (89.8) | 680 (85.8) |  | 915 (86.5) | 171 (85.1) | 744 (86.8) |
| Yes | 122 (13.4) | 12 (10.2) | 110 (13.8) |  | 139 (13.1) | 29 (14.4) | 110 (12.8) |
| No answer | 3 (0.3) | 0 (0.0) | 3 (0.4) |  | 4 (0.4) | 1 (0.5) | 3 (0.4) |
| Self-reported diabetes |  |  |  |  |  |  |  |
| No | 803 (88.1) | 109 (92.4) | 694 (87.5) |  | 934 (88.3) | 172 (85.6) | 762 (88.9) |
| Yes | 105 (11.5) | 9 (7.6) | 96 (12.1) |  | 120 (11.3) | 28 (13.9) | 92 (10.7) |
| No answer | 3 (0.3) | 0 (0.0) | 3 (0.4) |  | 4 (0.4) | 1 (0.5) | 3 (0.4) |

| **eTable 2.** Household and community risk factors for TMA positivity and seropositivity among farmworkers, Monterey County, 2020. | | | | | | | |
| --- | --- | --- | --- | --- | --- | --- | --- |
| **Attribute** | **Current SARS-CoV-2 infection (TMA)** | | |  | **Historical SARS-CoV-2 infection (IgG reactivity)** | | |
|  | All enrolled  N=911 | Positive  N=118 | Negative  N=793 |  | All enrolled  *N*=1,058 | Positive  N=201 | Negative  N=857 |
|  | *n* (%) or M±SD | *n* (%) or M±SD | *n* (%) or M±SD |  | *n* (%) or M±SD | *n* (%) or M±SD | *n* (%) or M±SD |
| Type of housing |  |  |  |  |  |  |  |
| House | 443 (48.6) | 60 (50.9) | 383 (48.3) |  | 490 (46.3) | 101 (50.3) | 389 (45.4) |
| Apartment | 372 (40.8) | 48 (40.7) | 324 (40.9) |  | 468 (44.2) | 85 (42.3) | 383 (44.7) |
| Hotel or motel | 36 (4.0) | 6 (5.1) | 30 (3.8) |  | 37 (3.5) | 5 (2.5) | 32 (3.7) |
| Trailer or mobile home | 39 (4.3) | 3 (2.5) | 36 (4.5) |  | 41 (3.9) | 8 (4.0) | 33 (3.9) |
| Other | 21 (2.3) | 1 (0.9) | 20 (2.5) |  | 22 (2.1) | 2 (1.0) | 20 (2.3) |
| Household size | 5.4 ± 2.3 |  |  |  | 5.5 ± 2.6 |  |  |
| 0 others | 11 (1.2) | 2 (1.7) | 9 (1.1) |  | 12 (1.1) | 3 (1.5) | 9 (1.1) |
| 1-3 others | 331 (36.3) | 41 (34.8) | 290 (36.6) |  | 379 (35.8) | 58 (28.9) | 321 (37.5) |
| 4-6 others | 414 (45.4) | 51 (43.2) | 363 (45.8) |  | 486 (45.9) | 93 (46.3) | 393 (45.9) |
| ≥7 others | 155 (17.0) | 24 (20.3) | 131 (16.5) |  | 181 (17.1) | 47 (23.4) | 134 (15.6) |
| Children <18 living in the home | 1.8 ± 1.5 | 1.8 ± 1.5 | 1.8 ± 1.6 |  | 1.8 ± 1.5 | 2.1 ± 1.7 | 1.7 ± 1.5 |
| No | 235 (25.8) | 28 (23.7) | 207 (26.1) |  | 268 (25.3) | 43 (21.4) | 225 (26.3) |
| Yes | 675 (74.1) | 90 (76.3) | 585 (73.8) |  | 789 (74.6) | 157 (78.1) | 632 (73.8) |
| No answer | 1 (0.1) | 0 (0.0) | 1 (0.1) |  | 1 (0.1) | 1 (0.5) | 0 (0.0) |
| Children ≤5 living in the home |  |  |  |  |  |  |  |
| No | 577 (63.3) | 79 (70.0) | 498 (62.8) |  | 669 (63.2) | 110 (54.7) | 559 (65.2) |
| Yes | 334 (36.7) | 39 (33.1) | 295 (37.2) |  | 389 (36.8) | 91 (45.3) | 298 (34.8) |
| Children attending school/daycare |  |  |  |  |  |  |  |
| No | 831 (91.2) | 105 (89.0) | 726 (91.6) |  | 976 (92.3) | 184 (91.5) | 792 (92.4) |
| Yes | 76 (8.3) | 12 (10.2) | 64 (8.1) |  | 78 (7.4) | 16 (8.0) | 62 (7.2) |
| No answer | 4 (0.4) | 1 (0.9) | 3 (0.4) |  | 4 (0.4) | 1 (0.5) | 3 (0.4) |
| Living with unrelated roommates |  |  |  |  |  |  |  |
| No | 732 (80.4) | 93 (78.8) | 639 (80.6) |  | 860 (81.3) | 156 (77.6) | 704 (82.2) |
| Yes | 179 (19.7) | 25 (21.2) | 154 (19.4) |  | 198 (18.7) | 45 (22.4) | 153 (17.9) |
| Living with other farmworkers | 1.6 ± 2.0 | 1.7 ± 1.6 | 1.6 ± 2.1 |  | 1.6 ± 1.8 | 1.6 ± 1.9 | 1.7 ± 1.7 |
| No | 224 (24.6) | 26 (22.0) | 198 (25.0) |  | 267 (25.2) | 49 (24.4) | 218 (25.4) |
| Yes | 684 (75.1) | 92 (78.0) | 592 (74.7) |  | 788 (74.5) | 151 (75.1) | 637 (74.3) |
| No answer | 3 (0.3) | 0 (0.0) | 3 (0.4) |  | 4 (0.3) | 1 (0.5) | 2 (0.2) |
| Household crowding |  |  |  |  |  |  |  |
| ≤2 persons per bedroom | 576 (63.2) | 71 (60.2) | 505 (63.7) |  | 666 (63.0) | 113 (56.2) | 553 (64.5) |
| >2 persons per bedroom | 335 (36.8) | 47 (39.8) | 288 (36.3) |  | 392 (37.1) | 88 (43.8) | 304 (35.5) |
| Access to washing machine at home |  |  |  |  |  |  |  |
| No | 345 (37.9) | 49 (41.5) | 296 (37.3) |  | 395 (37.3) | 77 (38.3) | 318 (37.1) |
| Yes | 566 (62.1) | 69 (58.5) | 497 (62.7) |  | 663 (62.7) | 124 (61.7) | 539 (62.9) |
| Left home for non-essential reasons (past 2 weeks) |  |  |  |  |  |  |  |
| No | 797 (87.5) | 104 (88.1) | 693 (87.4) |  | 914 (86.4) | 169 (84.1) | 745 (86.9) |
| Yes | 109 (12.0) | 12 (10.2) | 97 (12.2) |  | 138 (13.0) | 29 (14.4) | 109 (12.7) |
| No answer | 5 (0.6) | 2 (1.7) | 3 (0.4) |  | 6 (0.6) | 3 (1.5) | 3 (0.4) |
| Used public transportation/ride share services (past 2 weeks) |  |  |  |  |  |  |  |
| No | 858 (94.2) | 113 (95.8) | 745 (94.0) |  | 990 (93.6) | 189 (94.0) | 801 (93.5) |
| Yes | 48 (5.3) | 3 (2.5) | 45 (5.7) |  | 62 (5.9) | 0 )4.5) | 53 (6.2) |
| No answer | 5 (0.6) | 2 (1.7) | 3 (0.4) |  | 6 (0.6) | 3 (1.5) | 3 (0.4) |
| Attended social gatherings with non-household members (past 2 weeks) |  |  |  |  |  |  |  |
| No | 819 (89.9) | 105 (89.0) | 714 (90.0) |  | 949 (89.7) | 181 (90.1) | 768 (89.6) |
| Yes | 90 (9.9) | 13 (11.0) | 77 (9.7) |  | 108 (10.2) | 20 (10.0) | 88 (10.3) |
| No answer | 2 (0.2) | 0 (0.0) | 2 (0.3) |  | 1 (0.1) | 0 (0.0) | 1 (0.1) |
| Attended indoor gatherings with non-household members (past 2 weeks) |  |  |  |  |  |  |  |
| No | 862 (94.6) | 109 (92.4) | 753 (95.0) |  | 999 (94.4) | 192 (95.5) | 807 (94.2) |
| Yes | 47 (5.2) | 9 (7.6) | 38 (4.8) |  | 58 (5.5) | 9 (4.5) | 49 (5.7) |
| No answer | 2 (0.2) | 0 (0.0) | 2 (0.3) |  | 1 (0.1) | 0 (0.0) | 1 (0.1) |
| Face covering use while less than 6 feet away from others all of the time |  |  |  |  |  |  |  |
| No | 74 (8.1) | 114 (96.6) | 723 (91.2) |  | 81 (7.7) | 188 (95.5) | 789 (92.1) |
| Yes | 837 (91.9) | 4 (3.4) | 70 (8.8) |  | 977 (92.3) | 13 (6.5) | 68 (7.9) |
| Hand washing when returning home or after touching something all or most of the time |  |  |  |  |  |  |  |
| No | 27 (3.0) | 3 (2.5) | 24 (3.0) |  | 31 (2.9) | 5 (2.5) | 26 (3.0) |
| Yes | 884 (97.0) | 115 (97.5) | 769 (97.0) |  | 1,027 (97.1) | 196 (97.5) | 831 (97.0) |
| Possible exposure to someone with COVID-19 at home in the preceding two weeks^a^ |  |  |  |  |  |  |  |
| No | 804 (88.3) | 79 (67.0) | 725 (91.4) |  | 945 (89.3) | 178 (88.6) | 767 (89.5) |
| Yes | 107 (11.8) | 39 (33.1) | 68 (8.6) |  | 113 (10.7) | 23 (11.4) | 90 (10.5) |
| Possible exposure to someone with COVID-19 at home since the start of the pandemic^b^ |  |  |  |  |  |  |  |
| No | 743 (81.6) | 68 (57.6) | 675 (85.1) |  | 875 (82.7) | 152 (75.6) | 723 (84.4) |
| Yes | 168 (18.4) | 50 (42.4) | 118 (14.9) |  | 183 (17.3) | 49 (24.4) | 134 (15.6) |
| ^a^Lived with someone who had COVID-19 symptoms or positive in the preceding two weeks.  ^b^Lived with someone who had COVID-19 symptoms or positive since the start of the pandemic. | | | | | | | |

| **eTable 3.** Work-related risk factors for TMA positivity and seropositivity among farmworkers, Monterey County, 2020. | | | | | | | |
| --- | --- | --- | --- | --- | --- | --- | --- |
| **Attribute** | **Current SARS-CoV-2 infection (TMA)** | | |  | **Historical SARS-CoV-2 infection (IgG reactivity)** | | |
|  | All enrolled  N=911 | Positive  N=118 | Negative  N=793 |  | All enrolled  N=1058 | Positive  N=201 | Negative  N=857 |
|  | *n* (%) or M±SD | *n* (%) or M±SD | *n* (%) or M±SD |  | *n* (%) or M±SD | *n* (%) or M±SD | *n* (%) or M±SD |
| H2A visa holder |  |  |  |  |  |  |  |
| No | 840 (92.2) | 107 (90.7) | 733 (92.4) |  | 980 (92.6) | 188 (93.5) | 792 (92.4) |
| Yes | 60 (6.6) | 9 (7.6) | 51 (6.4) |  | 65 (6.1) | 11 (5.5) | 54 (6.3) |
| No answer | 11 (1.2) | 2 (1.7) | 9 (1.1) |  | 13 (1.2) | 2 (1.0) | 11 (1.3) |
| Supervisor or mayordomo |  |  |  |  |  |  |  |
| No | 867 (95.2) | 111 (94.1) | 756 (95.3) |  | 967 (91.4) | 183 (91.0) | 784 (91.5) |
| Yes | 44 (4.8) | 7 (5.9) | 37 (4.7) |  | 49 (4.6) | 9 (4.5) | 40 (4.7) |
| No answer | 0 (0.0) | 0 (0.0) | 0 (0.0) |  | 42 (4.0) | 9 (4.5) | 33 (3.9) |
| Type of agricultural work (ever/in the preceding two weeks) |  |  |  |  |  |  |  |
| Working in the fields | 680 (74.6) | 100 (84.8) | 580 (73.1) |  | 795 (75.1) | 162 (80.6) | 633 (73.9) |
| Packing shed | 105 (11.5) | 11 (9.3) | 94 (11.9) |  | 128 (12.1) | 21 (10.5) | 107 (12.5) |
| Processing facility | 57 (6.3) | 4 (3.4) | 53 (6.7) |  | 58 (5.5) | 7 (3.5) | 51 (6.0) |
| Nursery | 33 (3.6) | 4 (3.4) | 29 (3.7) |  | 35 (3.3) | 4 (2.0) | 31 (3.6) |
| Truck driver | 33 (3.6) | 4 (3.4) | 29 (3.7 |  | 33 (3.1) | 3 (1.5) | 30 (3.5) |
| Packing truck | 21 (2.3) | 1 (0.9) | 20 (2.5) |  | 21 (2.0) | 2 (1.0) | 8 (0.9) |
| Other | 19 (2.1) | 1 (0.9) | 18 (2.3) |  | 19 (1.8) | 2 (1.0) | 17 (2.0) |
| No answer | 1 (0.1) | 0 (0.0) | 1 (0.1) |  | 10 (0.1) | 2 (1.0) | 8 (0.9) |
| Worked indoors |  |  |  |  |  |  |  |
| No | 687 (75.5) | 98 (93.1) | 589 (74.3) |  | 812 (76.8) | 166 (82.6) | 646 (65.4) |
| Yes | 223 (24.5) | 20 (17.0) | 203 (25.6) |  | 245 (23.2) | 35 (17.4) | 210 (24.5) |
| No answer | 1 (0.1) | 0 (0.0) | 1 (0.1) |  | 1 (0.1) | 0 (0.0) | 1 (0.1) |
| Crops (ever/in the preceding two weeks) |  |  |  |  |  |  |  |
| Berries | 195 (28.7) | 14 (14.0) | 181 (31.2) |  | 233 (29.3) | 39 (24.1) | 194 (30.7) |
| Leafy greens | 173 (25.4) | 31 (31.0) | 142 (24.5) |  | 207 (26.0) | 50 (30.9) | 157 (24.8) |
| Broccoli | 148 (21.8) | 28 (28.0) | 120 (20.7) |  | 147 (18.5) | 24 (14.8) | 123 (19.4) |
| Grapes | 47 (6.9) | 10 (10.0) | 37 (6.4) |  | 57 (7.2) | 15 (9.3) | 42 (6.6) |
| Peas | 52 (7.7) | 16 (16.0) | 36 (6.2) |  | 50 (6.3) | 5 (3.1) | 45 (7.1) |
| Cauliflower | 37 (5.4) | 5 (5.0) | 32 (5.5) |  | 36 (4.5) | 11 (6.8) | 25 (4.0) |
| Celery | 17 (2.5) | 2 (2.0) | 15 (2.6) |  | 16 (2.0) | 3 (1.9) | 13 (2.1) |
| Artichokes | 5 (0.7) | 0 (0.0) | 5 (0.9) |  | 6 (0.8) | 1 (0.6) | 5 (0.8) |
| Other | 107 (15.7) | 9 (9.0) | 98 (16.9) |  | 155 (19.5) | 35 (21.6) | 120 (19.0) |
| Commuted to work with non-household members |  |  |  |  |  |  |  |
| No | 595 (65.3) | 65 (55.1) | 530 (66.8) |  | 672 (63.5) | 124 (61.7) | 548 (63.9) |
| Yes | 316 (34.7) | 53 (44.9) | 263 (33.2) |  | 366 (34.6) | 72 (35.8) | 294 (34.3) |
| No answer | 0 (0.0) | 0 (0.0) | 0 (0.0) |  | 20 (1.9) | 5 (2.5) | 15 (1.8) |
| Used face covering at work all of the time |  |  |  |  |  |  |  |
| No | 94 (10.3) | 11 (9.3) | 83 (10.5) |  | 105 (9.9) | 19 (9.5) | 86 (10.0) |
| Yes | 816 (89.7) | 107 (90.7) | 709 (89.4) |  | 950 (89.8) | 182 (90.1) | 768 (89.6) |
| No answer | 1 (0.1) | 0 (0.0) | 1 (0.1) |  | 3 (0.3) | 0 (0.0) | 3 (0.4) |
| Came within 6 feet from others while working |  |  |  |  |  |  |  |
| No | 413 (45.3) | 52 (44.1) | 361 (45.5) |  | 483 (45.7) | 95 (47.3) | 388 (45.3) |
| Yes | 477 (52.4) | 64 (54.2) | 413 (52.1) |  | 538 (50.1) | 97 (48.3) | 441 (51.5) |
| No answer | 21 (2.3) | 2 (1.7) | 19 (2.4) |  | 37 (4.2) | 9 (4.5) | 28 (23.3) |
| Possible exposure to someone with COVID-19 at work in the preceding two weeks^a^ |  |  |  |  |  |  |  |
| No | 763 (83.8) | 83 (70.3) | 680 ((85.8) |  | 922 (87.2) | 178 (88.6) | 744 (86.8) |
| Yes | 148 (16.3) | 35 (29.7) | 113 (14.3) |  | 136 (12.9) | 23 (11.4) | 113 (13.2) |
| No answer | 0 (0.0) | 0 (0.0) | 0 (0.0) |  | 0 (0.0) |  |  |
| Possible exposure to someone with COVID-19 at work since the start of the pandemic^b^ |  |  |  |  |  |  |  |
| No | 523 (57.4) | 52 (44.1) | 471 (59.4) |  | 641 (60.6) | 117 (58.2) | 524 (61.1) |
| Yes | 388 (42.6) | 66 (55.9) | 322 (40.6) |  | 417 (39.4) | 84 (41.8) | 333 (38.9) |
| No answer | 0 (0.0) | 0 (0.0) | 0 (0.0) |  | 0 (0.0) | 0 (0.0) | 0 (0.0) |
| ^a^Worked with someone who had COVID-19 symptoms, tested positive for SARS-CoV-2, or who quarantined in the preceding two weeks.  ^b^Worked with someone who had COVID-19 symptoms, tested positive for SARS-CoV-2, or who quarantined since the start of the pandemic. | | | | | | | |

| **eTable 4.** Employer-provided preventive measures and their association with TMA positivity and seropositivity among farmworkers, Monterey County, 2020. | | | | | | | |
| --- | --- | --- | --- | --- | --- | --- | --- |
| **Attribute** | **Current SARS-CoV-2 infection (TMA)** | | |  | **Historical SARS-CoV-2 infection (IgG reactivity)** | | |
|  | All enrolled  N=911 | Positive  N=118 | Negative  N=793 |  | All enrolled  N=1058 | Positive  N=201 | Negative  N=857 |
|  | *n* (%) or M±SD | *n* (%) or M±SD | *n* (%) or M±SD |  | *n* (%) or M±SD | *n* (%) or M±SD | *n* (%) or M±SD |
| Fever and symptoms screening upon arrival at workplace |  |  |  |  |  |  |  |
| Neither | 326 (35.8) | 54 (45.8) | 272 (34.3) |  | 480 (45.4) | 92 (45.8) | 388 (45.3) |
| Either or both | 585 (64.2) | 64 (54.2) | 521 (65.7) |  | 577 (54.5) | 109 (54.2) | 468 (54.6) |
| No answer | 0 (0.0) | 0 (0.0) | 0 (0.0) |  | 1 (0.1) | 0 (0.0) | 1 (0.1) |
| Employer provided face coverings |  |  |  |  |  |  |  |
| No | 494 (54.2) | 15 (12.7) | 109 (13.8) |  | 162 (15.3) | 26 (12.9) | 136 (15.9) |
| Yes | 413 (45.3) | 102 (86.4) | 681 (85.9) |  | 889 (84.0) | 174 (86.6) | 715 (83.4) |
| No answer | 4 (0.4) | 1 (0.9) | 3 (0.4) |  | 7 (0.7) | 1 (0.5) | 6 (0.7) |
| Employer provided gloves |  |  |  |  |  |  |  |
| No | 133 (14.6) | 12 (10.2) | 121 (15.3) |  | 152 (14.4) | 26 (12.9) | 126 (14.7) |
| Yes | 777 (85.3) | 106 (89.8) | 671 (84.6) |  | 905 (85.5) | 175 (87.1) | 730 (85.2) |
| No answer | 1 (0.1) | 0 (0.0) | 1 (0.1) |  | 1 (0.1) | 0 (0.0) | 1 (0.1) |
| Employer provided eye shields |  |  |  |  |  |  |  |
| No | 444 (48.7) | 52 (44.1) | 392 (49.4) |  | 518 (49.0) | 94 (46.8) | 424 (49.5) |
| Yes | 466 (51.2) | 66 (55.9) | 400 (50.4) |  | 539 (50.1) | 107 (53.2) | 432 (50.4) |
| No answer | 1 (0.1) | 0 (0.0) | 1 (0.1) |  | 1 (0.1) | 0 (0.0) | 1 (0.1) |
| Employer provided hand washing stations |  |  |  |  |  |  |  |
| No | 5 (0.6) | 1 (0.9) | 4 (0.5) |  | 6 (0.6) | 2 (1.0) | 4 (0.5) |
| Yes | 905 (99.3) | 117 (99.2) | 788 (99.4) |  | 1,051 (99.3) | 199 (99.0) | 852 (99.4) |
| No answer | 1 (0.1) | 0 (0.0) | 1 (0.1) |  | 1 (0.1) | 0 (0.0) | 1 (0.1) |
| Employer provided liquid soap and paper towels |  |  |  |  |  |  |  |
| No | 12 (1.3) | 2 (1.7) | 10 (1.3) |  | 15 (1.4) | 4 (2.0) | 11 (1.3) |
| Yes | 897 (98.5) | 116 (98.3) | 781 (98.5) |  | 1,041 (98.4) | 196 (97.5) | 845 (98.6) |
| No answer | 2 (0.2) | 0 (0.0) | 2 (0.3) |  | 2 (0.2) | 1 (0.5) | 1 (0.1) |
| Employer provided hand sanitizer |  |  |  |  |  |  |  |
| No | 70 (7.7) | 8 (6.8) | 62 (7.8) |  | 89 (8.4) | 20 (10.0) | 69 (9.1) |
| Yes | 840 (92.2) | 110 (93.2) | 730 (92.1) |  | 968 (91.5) | 181 (90.1) | 787 (91.8) |
| No answer | 1 (0.1) | 0 (0.0) | 1 (0.1) |  | 1 (0.1) | 0 (0.0) | 1 (0.1) |
| Workplace surfaces and tools regularly disinfected and kept clean |  |  |  |  |  |  |  |
| No | 94 (10.3) | 12 (10.2) | 82 (10.3) |  | 115 (10.9) | 17 (8.5) | 98 (11.4) |
| Yes | 785 (86.2) | 98 (83.1) | 687 (86.6) |  | 905 (85.5) | 175 (87.1) | 730 (85.2) |
| No answer | 32 (3.5) | 8 (6.8) | 23 (3.0) |  | 38 (3.6) | 9 (4.5) | 29 (3.4) |
| Employer staggered breaks to reduce exposure |  |  |  |  |  |  |  |
| No | 494 (54.2) | 66 (55.9) | 428 (54.0) |  | 583 (55.1) | 112 (55.7) | 471 (55.0) |
| Yes | 413 (45.3) | 51 (43.2) | 362 (45.7) |  | 469 (44.3) | 89 (44.3) | 380 (44.3) |
| No answer | 4 (0.4) | 1 (0.9) | 3 (0.4) |  | 6 (0.6) | 0 (0.0) | 6 (0.7) |
| Employer provided information on COVID-19 symptoms |  |  |  |  |  |  |  |
| No | 40 (4.4) | 6 (5.1) | 34 (4.3) |  | 62 (5.9) | 17 (8.5) | 45 (5.3) |
| Yes | 870 (95.5) | 112 (94.9) | 758 (95.6) |  | 993 (93.9) | 184 (91.5) | 809 (94.4) |
| No answer | 1 (0.1) | 0 (0.0) | 1 (0.1) |  | 3 (0.3) | 0 (0.0) | 3 (0.4) |
| Employer provided information on how to protect themselves at work |  |  |  |  |  |  |  |
| No | 21 (2.3) | 1 (0.9) | 20 (2.5) |  | 35 (3.3) | 12 (6.0) | 23 (2.7) |
| Yes | 889 (97.6) | 117 (99.2) | 772 (97.4) |  | 1,020 (96.4) | 189 (94.0) | 831 (97.0) |
| No answer | 1 (0.11) | 0 (0.0) | 1 (0.1) |  | 3 (0.3) | 0 (0.0) | 3 (0.4) |
| Employer provided information on how to protect themselves at home and in the community |  |  |  |  |  |  |  |
| No | 47 (5.2) | 3 (2.5) | 44 (5.6) |  | 67 (6.3) | 17 (8.5) | 50 (5.8) |
| Yes | 863 (94.7) | 115 (97.5) | 748 (94.3) |  | 988 (93.4) | 184 (91.5) | 804 (93.8) |
| No answer | 1 (0.11) | 0 (0.0) | 1 (0.1) |  | 3 (0.3) | 0 (0.0) | 3 (0.4) |
| Employer provided information on whom to call if they were sick |  |  |  |  |  |  |  |
| No | 99 (10.9) | 18 (15.3) | 81 (10.2) |  | 129 (12.2) | 27 (13.4) | 102 (11.9) |
| Yes | 811 (89.0) | 100 (84.8) | 711 (89.7) |  | 926 (87.5) | 174 (86.6) | 752 (87.8) |
| No answer | 1 (0.11) | 0 (0.0) | 1 (0.1) |  | 3 (0.3) | 0 (0.0) | 3 (0.4) |
| Employer provided information on their ability to get free testing and treatment if they were sick |  |  |  |  |  |  |  |
| No | 237 (26.0) | 31 (26.3) | 206 (26.0) |  | 289 (27.3) | 62 (30.9) | 227 (26.5) |
| Yes | 672 (73.8) | 87 (73.7) | 585 (73.8) |  | 765 (72.3) | 138 (68.7) | 627 (73.2) |
| No answer | 2 (0.2) | 0 (0.0) | 2 (0.3) |  | 4 (0.4) | 1 (0.5) | 3 (0.4) |
| Employer provided information on where to get housing if they needed to quarantine or isolate away from home |  |  |  |  |  |  |  |
| No | 498 (54.7) | 70 (59.3) | 428 (54.0) |  | 579 (54.7) | 112 (55.7) | 467 (54.5) |
| Yes | 411 (45.1) | 48 (40.7) | 363 (45.8) |  | 475 (44.9) | 88 (43.8) | 387 (45.2) |
| No answer | 2 (0.22) | 0 (0.0) | 2 (0.3) |  | 4 (0.4) | 1 (0.5) | 3 (0.4) |
| Employer provided information on the importance of staying away from work if they were sick |  |  |  |  |  |  |  |
| No | 55 (6.0) | 5 (4.2) | 50 (6.3) |  | 77 (7.3) | 15 (7.5) | 62 (7.2) |
| Yes | 854 (93.7) | 113 (96.0) | 741 (93.4) |  | 976 (92.3) | 185 (92.0) | 791 (92.3) |
| No answer | 2 (0.2) | 0 (0.0) | 2 (0.3) |  | 5 (0.5) | 1 (0.5) | 4 (0.5) |
| Employer provided information on their benefit to get paid to stay away from work if they were sick |  |  |  |  |  |  |  |
| No | 263 (28.9) | 36 (30.5) | 227 (28.6) |  | 316 (29.9) | 58 (28.9) | 258 (30.1) |
| Yes | 646 (70.9) | 82 (69.5) | 564 (71.1) |  | 738 (69.8) | 142 (70.7) | 596 (69.5) |
| No answer | 2 (0.2) | 0 (0.0) | 2 (0.3) |  | 4 (0.4) | 1 (0.5) | 3 (0.4) |
| Received education about COVID-19 from medical staff at workplace |  |  |  |  |  |  |  |
| No | 584 (64.1) | 76 (64.4) | 508 (64.1) |  | 682 (64.5) | 126 (62.7) | 556 (64.9) |
| Yes | 312 (34.3) | 41 (34.8) | 271 (34.2) |  | 356 (33.7) | 73 (36.3) | 283 (33.0) |
| No answer | 15 (1.7) | 1 (0.9) | 13 (1.8) |  | 20 (1.9) | 2 (1.0) | 18 (2.1) |

**eFigure 1.** Correlation heat map of risk factors related to TMA positivity or seropositivity among farmworkers, Monterey County, 2020 (n=1107).

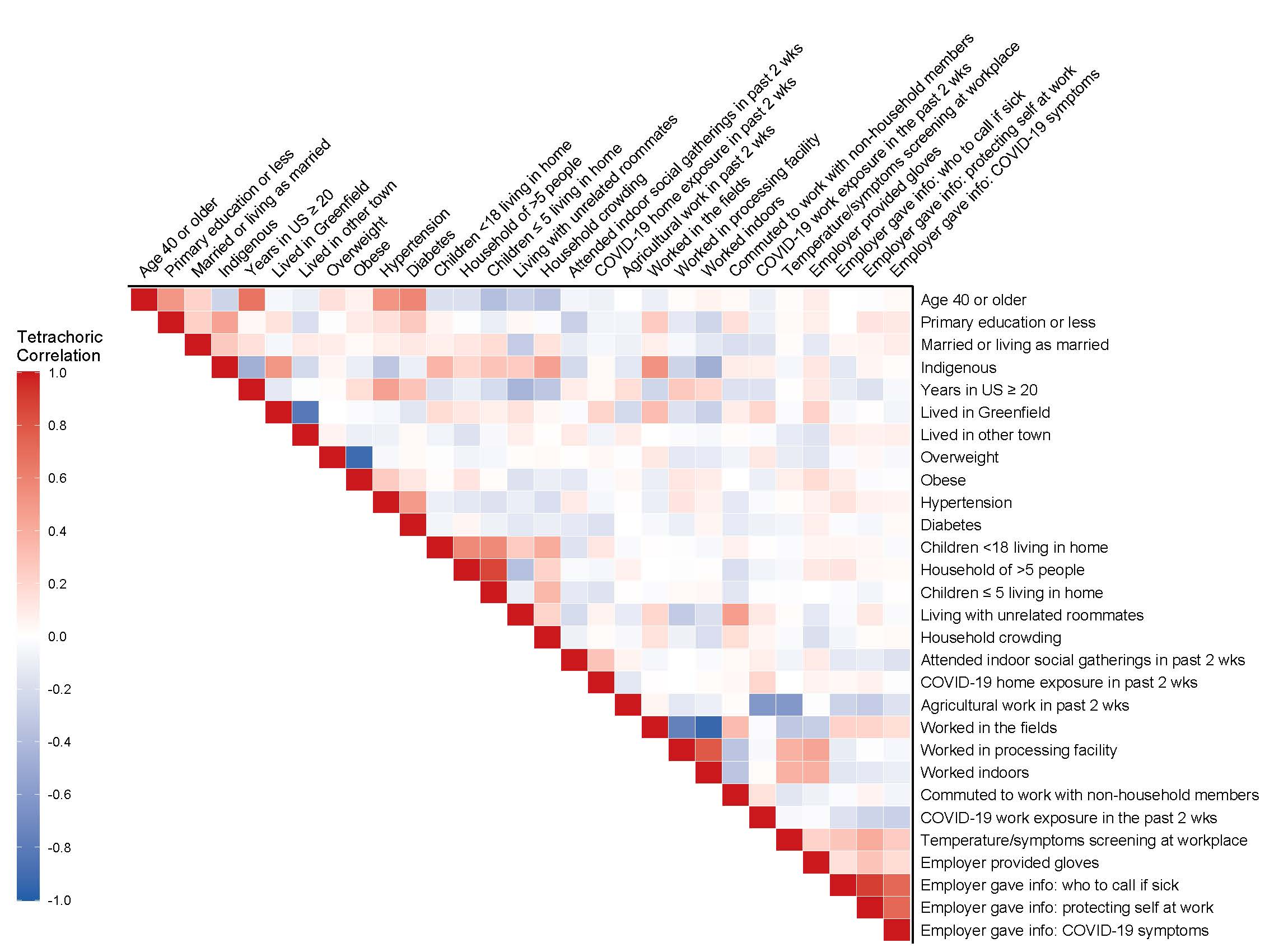
